# Insights from a nationwide study of 10,929 Australians living with Parkinson’s disease: Risk factors, comorbidities, and sex differences

**DOI:** 10.1101/2025.05.23.25328193

**Authors:** Fangyuan Cao, Kerrie McAloney, Natalia S. Ogonowski, Luis M. García-Marín, Santiago Díaz-Torres, Victor Flores-Ocampo, Zuriel Ceja, Freddy Chafota, Richard Parker, Mary Ferguson, Rebekah A. Cicero, Susan E. List-Armitage, Vicki Miller, Clyde Campbell, Carolyn M. Sue, Kishore R. Kumar, George D. Mellick, Nicholas G. Martin, Miguel E. Rentería

## Abstract

**Background:** Parkinson’s disease (PD) is a complex neurodegenerative condition with a heterogeneous clinical presentation and multifactorial aetiology. Despite substantial progress in PD research, the underlying causes and biological pathways of PD remain incompletely understood, warranting the need for large-scale studies to further elucidate genetic and environmental contributors. The Australian Parkinson’s Genetics Study (APGS) is an ongoing nationwide, population-based initiative established to advance understanding of the determinants and progression of PD.

**Methods:** We present a cross-sectional characterisation of 10,929 participants with self-reported PD recruited across Australia through a combination of assisted mail-outs, media outreach, and digital engagement. Participants complete comprehensive questionnaires capturing sociodemographic, clinical, environmental, lifestyle, and behavioural data, and provide saliva samples for genetic analysis. A control cohort is currently being recruited and is therefore not reported here.

**Findings:** The cohort is 63% male, with a mean age of 71 years and symptom onset at 64 years. Most participants report being diagnosed by a neurologist (79%), and 25% have a family history of PD. Non-motor symptoms such as depression, anxiety, and sleep disturbances are common, and sex-based differences were observed across clinical features, comorbidities, environmental exposures, and impulse control behaviours. Previously reported risk factors were notable in the cohort, including pesticide exposure (36%), traumatic brain injury (16%), and employment in high-risk industries (33%).

**Interpretation:** APGS represents the largest PD cohort in Australia and the largest active PD cohort globally. Its comprehensive design and ongoing expansion, including digital phenotyping and genomic profiling, position it as a transformative platform to inform risk prediction, biomarker discovery, and therapeutic development for PD. The recruitment success highlights the effectiveness of our innovative and cost-effective outreach strategies and the feasibility of large-scale, remote recruitment through government-supported mail-outs and media engagement.

**Funding:** APGS is supported by the Shake It Up Australia Foundation and the Michael J. Fox Foundation for Parkinson’s Research (MJFF-021952).

## Introduction

Parkinson’s disease (PD) is the second most prevalent neurological condition and a growing global health priority. It affects over 10 million people worldwide, a figure projected to at least double by 2050 due to population aging.^1^ In Australia, more than 150,000 people live with PD, with approximately 50 new diagnoses daily and an annual economic burden estimated at AUD 10 billion.^2^

While PD is primarily recognised as a movement disorder characterised by bradykinesia, rigidity, and tremor, a myriad of non-motor symptoms, including impaired olfaction, autonomic dysfunction, sleep disturbances, mood and anxiety disorders, cognitive decline, speech and language impairments, and pain also emerge, many of which often preceding motor signs or diagnosis by years. ^1,3–5^

Ongoing efforts to better understand and treat PD have led to notable progress, with 136 clinical trials active as of January 2024; however, few have progressed to Phase 3, particularly those targeting disease modification.^6^ Current treatments, such as levodopa and deep brain stimulation, improve symptom control but can cause complications over time,^1,7^ and no therapy currently halts or reverses the neurodegeneration. Advancing effective therapies requires deeper insight into the underlying causes and biological pathways of PD.

PD arises from a complex interplay of age, environmental, and genetic factors. Advancing age, especially beyond 60 years, remains the strongest risk factor.^7–9^ Other factors, including male sex, head trauma, high alcohol consumption, farming occupations, and exposure to pesticides, heavy metals, and environmental pollutants have also been linked to increased PD risk,^7,8,10^ though findings have not always been consistent.

Genetic factors play a crucial role, with twin heritability estimates at around 34%.^11^ Approximately 10-15% of cases involve rare, highly penetrant monogenic variants, while most are complex, shaped by multiple common genetic variants of small effect interacting with environmental exposures.^10,12,13^ Notably, genes implicated in monogenic PD, including *SNCA*, *LRRK2*, and *GBA*, also contribute to sporadic PD risk.^12^ Genome-wide association studies (GWAS) have further expanded our understanding of PD, identifying up to 134 independent risk variants.^14^ However, these loci explain only a small fraction of the observed heritability, highlighting substantial “missing heritability” that will require nearly tripling GWAS sample sizes to uncover.^13^

The Global Parkinson’s Genetics Program (GP2) is a major international initiative aimed at uncovering the genetic architecture of PD by harmonising data across diverse populations.^15^ Within this framework, the Australian Parkinson’s Genetics Study (APGS) is the largest ongoing national PD study and a transformative effort advance PD research. APGS aims to expand sample collection and genotyping by recruiting and following a nationwide cohort of 10,000 affected individuals and 10,000 unaffected controls. Having reached its target enrolment of cases, this manuscript presents a comprehensive characterisation of the APGS PD cohort to date, addressing sociodemographic, clinical, and lifestyle features, as well as sex-specific differences across these domains.

## Methods

### Study design

The APGS is a large-scale research initiative led by the QIMR Berghofer Medical Research Institute in Brisbane, Australia. It seeks to characterise the phenotypic, genetic, and environmental diversity of PD to identify factors influencing disease onset and progression, thereby enhancing our understanding of PD, informing screening, diagnostic, and prevention strategies. Several sub-studies have been launched within the APGS framework to further strengthen its translational potential for future discovery and intervention (see Supplementary Methods).

The study protocol was approved by the Human Research Ethics Committee of QIMR Berghofer (P3711). All participants provided informed consent, including agreement for future recontact. Data are securely managed in accordance with the Commonwealth Privacy Act (1988) and the National Health and Medical Research Council (NHMRC) Guidelines.

### Participants

The APGS collaborates with *Services Australia* (formerly the Australian Government Department of Human Services) to deliver an assisted mail-out, enabling outreach to a random sample of individuals across Australia prescribed PD-related medications listed under the Pharmaceutical Benefits Scheme (PBS, https://www.pbs.gov.au/pbs/). Recipient’s personal data remain secure and confidential; no identifiable information is shared with researchers.

In 2020, we conducted a pilot study,^16^ in which over 1,500 individuals were recruited and provided a saliva sample for DNA extraction. Building on this initial success and supported by GP2, the Shake It Up Australia Foundation, and the Michael J. Fox Foundation, we launched full-scale nationwide recruitment in April 2022, utilising a combination of assisted mail-outs, national communications campaign, and public appeal.

Eligibility for cases required being an Australian citizen or permanent resident, aged 40-75 years, with at least one PBS claim for PD-related medications within the past two years (see Supplementary Methods for detailed eligibility, medication list, and recruitment waves). Informed consent was obtained electronically prior to questionnaire completion.

### Procedures

After providing informed consent, participants completed a questionnaire, either online via the Qualtrics Platform (https://www.qualtrics.com) or by mail using a paper version. The questionnaire collects comprehensive information across multiple domains (Table 1), including sociodemographic characteristics, clinical features, lifestyle factors, environmental exposures, and medical and behavioural history. Eligible participants were then invited to provide a saliva sample for genetic analysis (see Supplementary Methods). More information is available on the study website: https://www.qimrb.edu.au/studies/apgs.

**Table 1.**
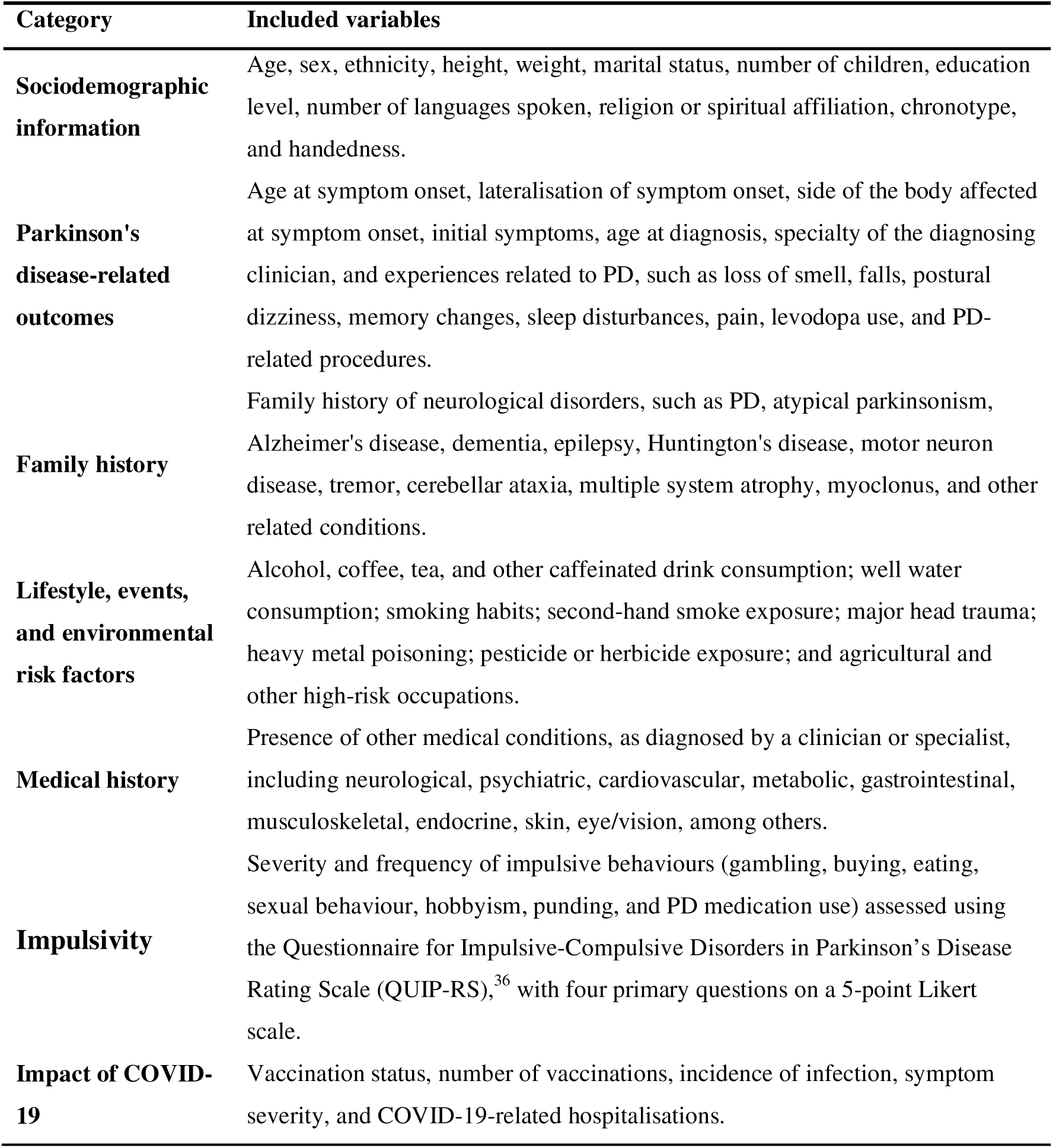
Overview of the APGS questionnaire.

### Statistical analysis

Baseline characteristics were reported as frequencies and percentages for categorical data and as means and standard deviations (SD) and medians and interquartile ranges (IQR) for continuous data. The number of participants available for each variable was also reported.

Independent t-tests and chi-square tests were performed to compare males and females on continuous and categorical variables, respectively. When continuous data deviated from normality, visually assessed by histograms and quantile-quantile (Q-Q) plots, the non-parametric Mann-Whitney U test was also applied. As results from parametric and non-parametric were nearly identical due to the large sample size, only parametric results are reported.

Statistical significance was set at α = 0.05. Analyses and visualisations were conducted in Python 3.12.5 using the *Pandas, NumPy, Scipy*, and *Matplotlib* libraries.

## Results

### Sample accrual

Recruitment occurred in phases between May 2020 and June 2025 (Figure 1a), with participant distribution by state and territory broadly mirroring the national population (Figure 1b). A pilot mail-out in mid-2020 prompted a sharp rise in participation, with over 1,000 questionnaires completed in June 2020. Monthly completions then declined but remained steady until a second peak in April 2021, following a national media campaign. The largest response came after a national mail-out of 100,000 letters in September 2022, peaking at over 1,200 completions in October. Participation tapered before rising again after a second mail-out in July 2023 (60,000 letters), which sustained moderate engagement into late 2023. A final mail-out in June 2024 (26,000 letters) led to another uptake in responses, enabling the study to reach its target of 10,000 participants diagnosed with PD.

**Figure 1.**
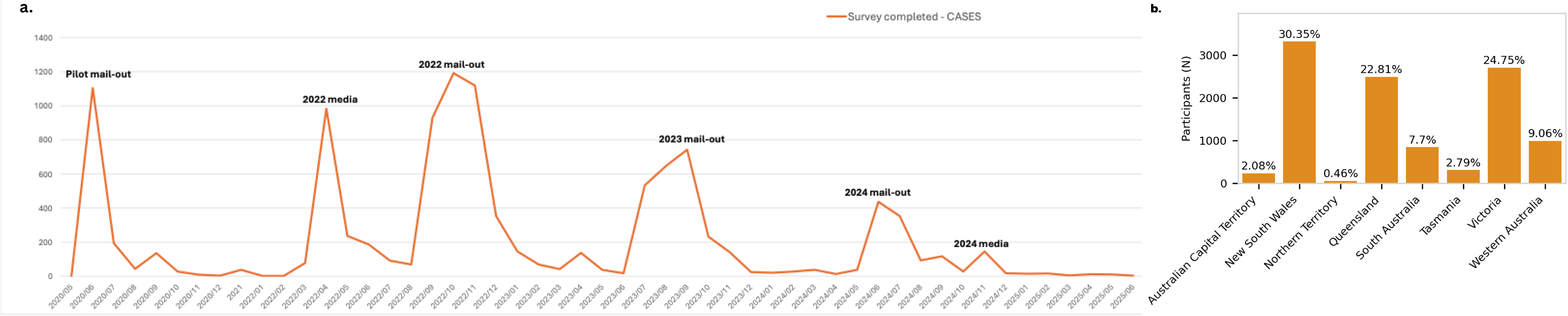
Temporal and geographic characteristics of APGS participant recruitment (May 2020-June 2025). **a.** illustrates the monthly number of surveys completed by APGS participants, showing patterns of engagement over time aligned with key recruitment phases. Peaks in survey completion correspond to major study outreach events, including the pilot mail-out (June 2020), national media campaigns in 2021 and 2024, and government-assisted invitation waves in 2022, 2023, and 2024. These recruitment efforts cumulatively enabled the enrolment of 10,000 individuals with Parkinson’s disease across Australia. **b.** shows the geographic composition of the APGS participants, which reflects the distribution of the Australian population.

### Demographics

Of the 10,929 participants with PD recruited by 12 June 2025, 88% responded via the online platform (Table 2). The cohort was 63% male, with an average age of 71 ± 9 years. The sample was predominantly of European ancestry (93%), non-religious (63%), married or in a de facto relationship (70%), and had children (89%), with 90% of those having two children or more. Overall, 73% completed at least high school, and 14% spoke more than one language. Most participants were right-handed (87%) and around half (51%) identified as “morning people.”

**Table 2.**
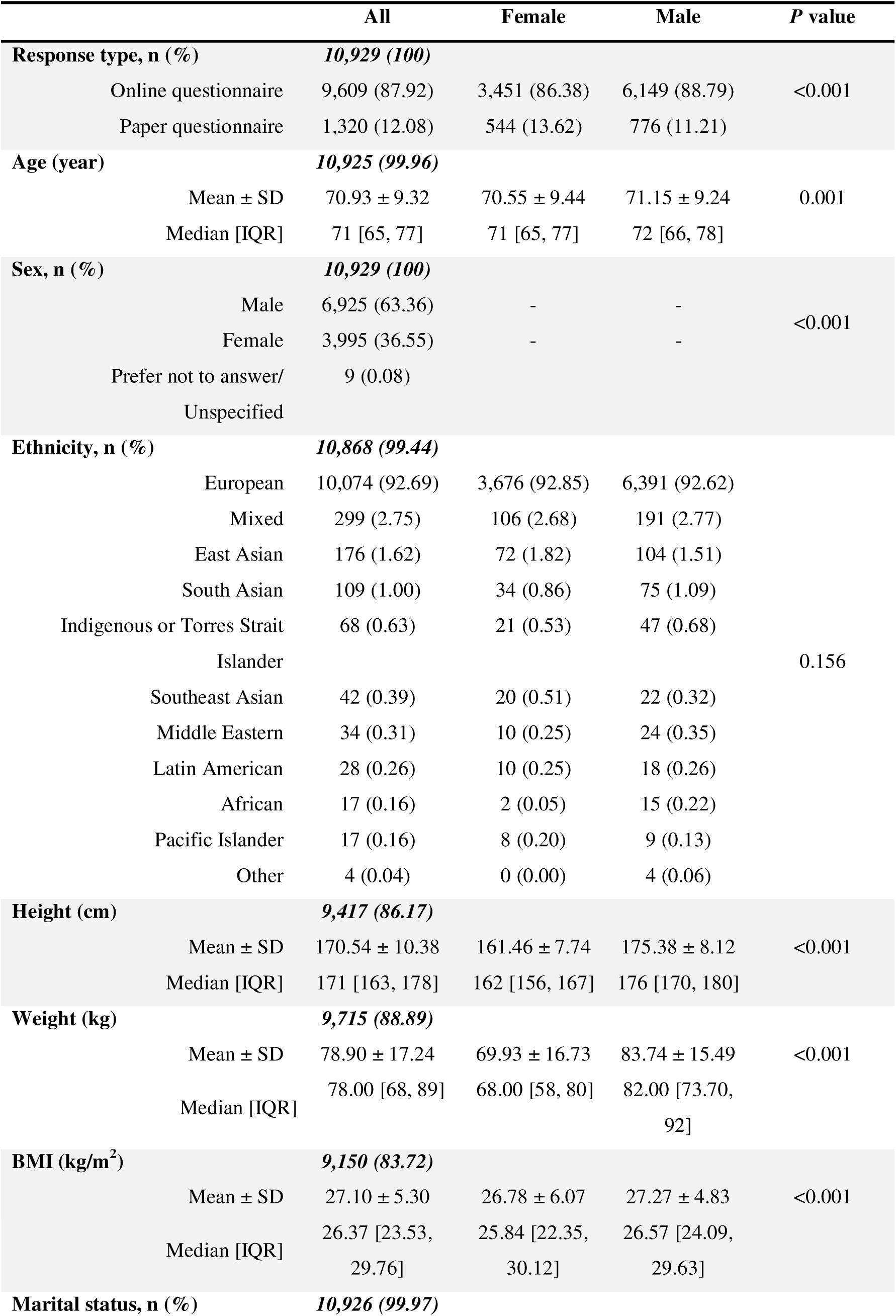
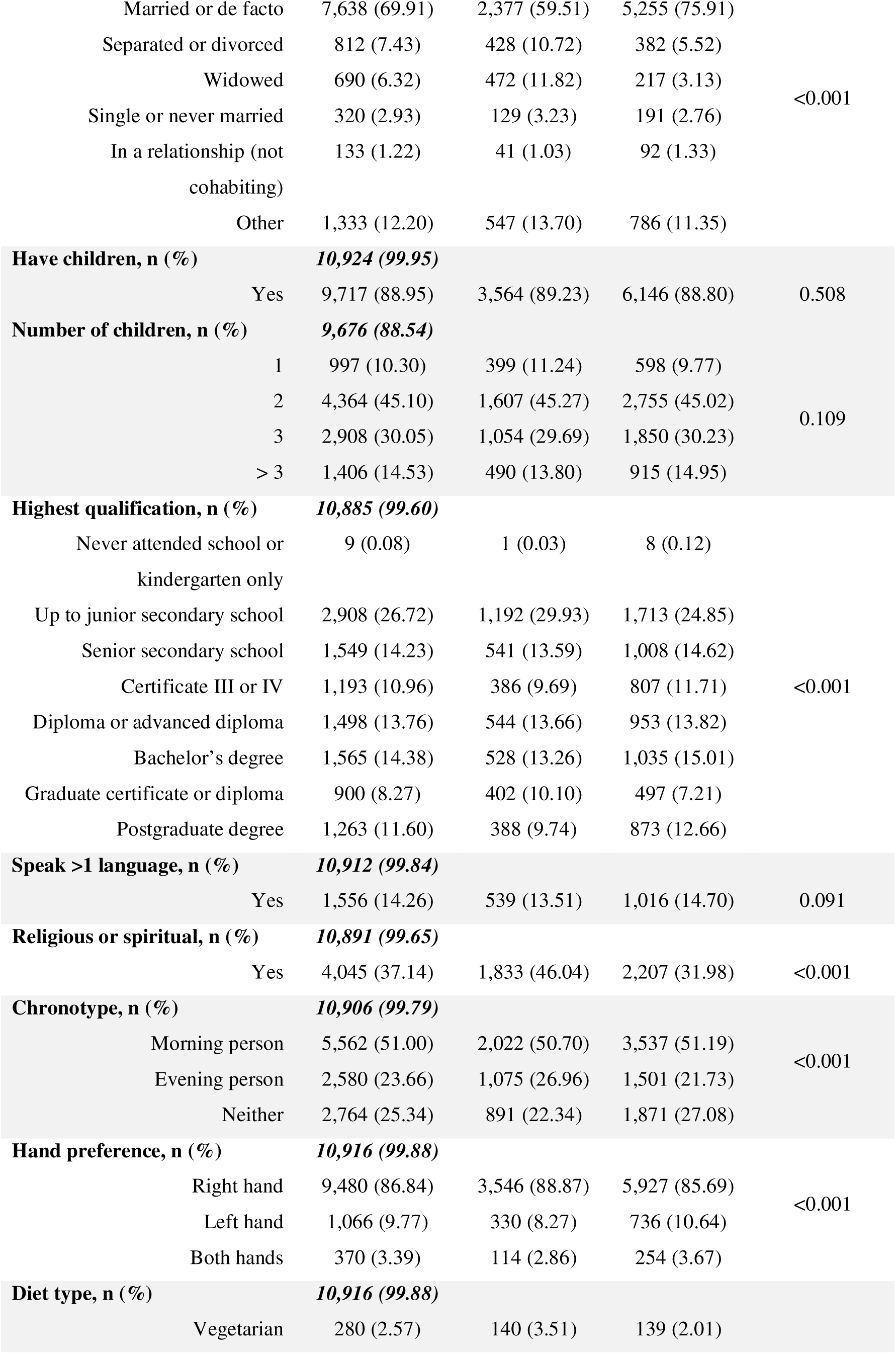
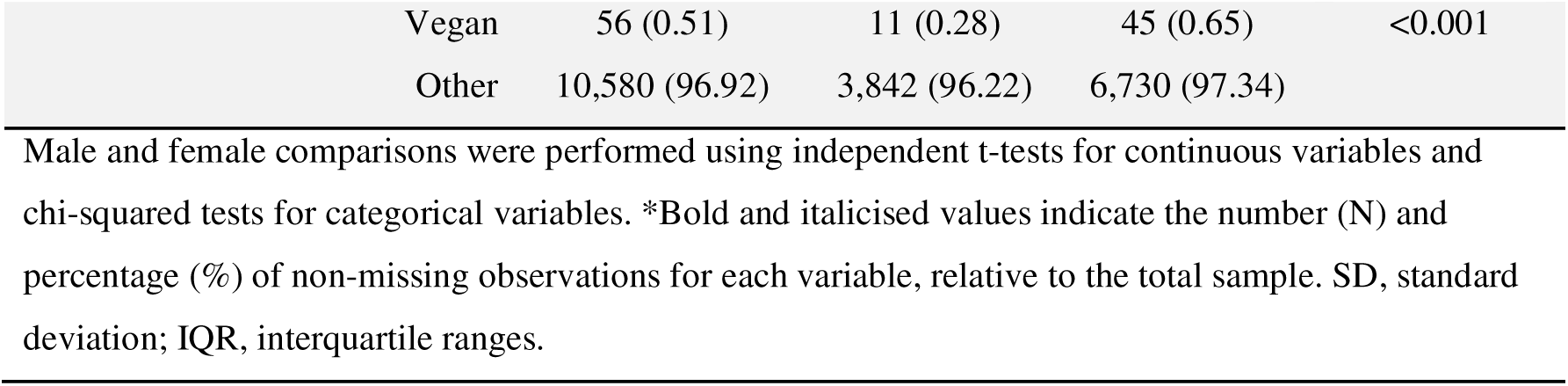
Sociodemographic characteristics of the APGS cohort.

Males and females had similar ages, with a difference of less than one year. Males were more likely to be married or in a de facto relationship (76% vs. 60%), left-handed (11% vs. 8%), or ambidextrous (4% vs. 3%), and had slightly higher education attainment (13% vs. 10% with a postgraduate degree). In contrast, females were more likely to be divorced or separated (11% vs. 6%), widowed (12% vs. 3%), religious or spiritual (46% vs. 32%), vegetarian (4% vs. 2%), and to identify as “evening people” (27% vs. 22%) (*p* < 0.001 for all comparisons).

### PD-related outcomes

Most participants (79%) reported being diagnosed by a neurologist, with a mean age of 64 ± 11 years at symptom onset and 68 ± 10 years at diagnosis (Table 3). Unilateral symptom onset was common (77%), with the right side affected in 53%. The most frequently reported initial symptom was tremor or shaking (52%), followed by bradykinesia (13%) and rigidity or stiffness (7%). Over half experienced concurrent symptoms such as hyposmia, orthostatic light-headedness, memory changes, and pain. Notably, 96% of participants reported sleep disturbances, including insomnia, daytime sleepiness, vivid dreams, dream enactment, or restless legs. In terms of treatment, 94% had been prescribed levodopa, and 6% had undergone neurosurgical intervention, primarily deep brain stimulation.

**Table 3.**
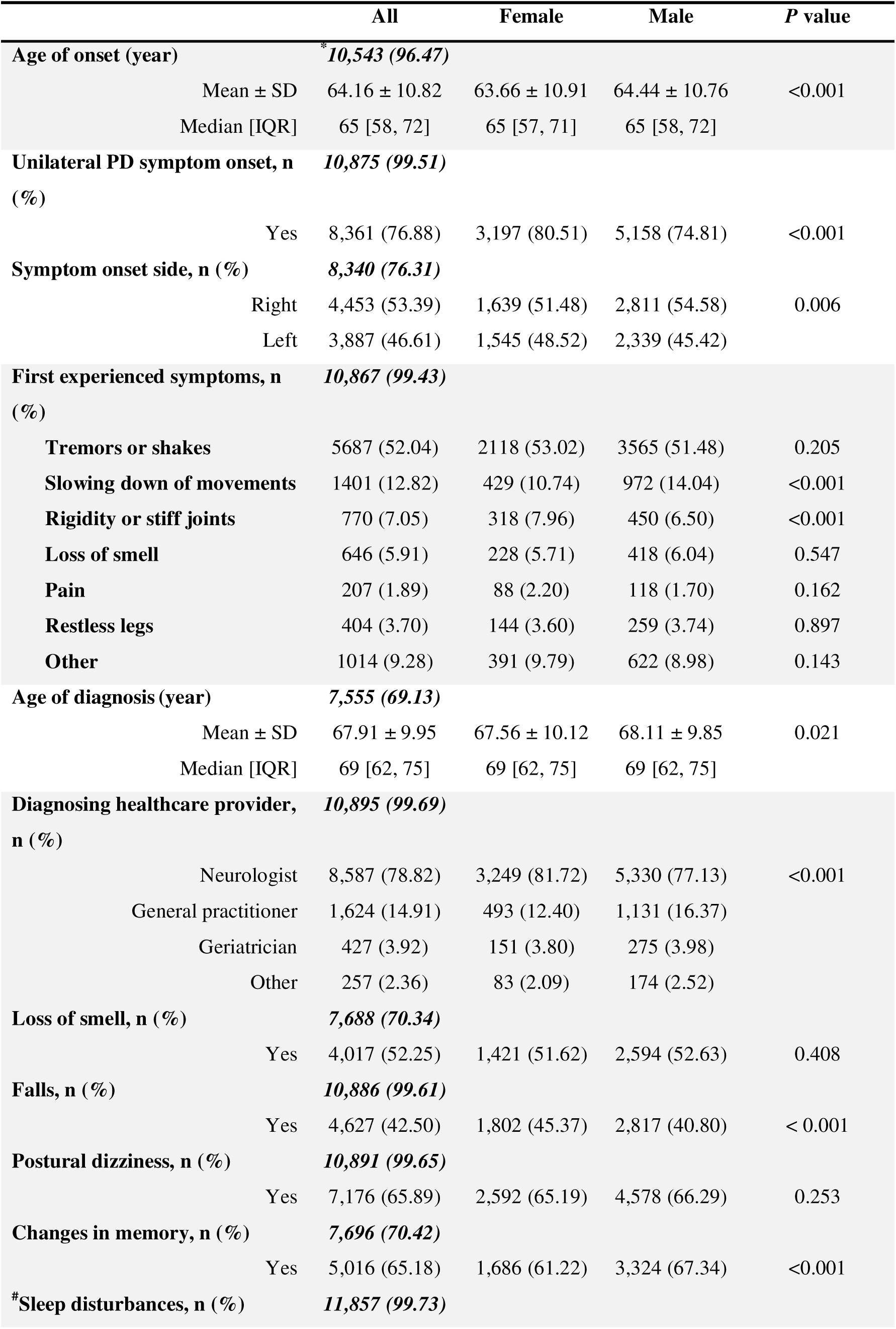
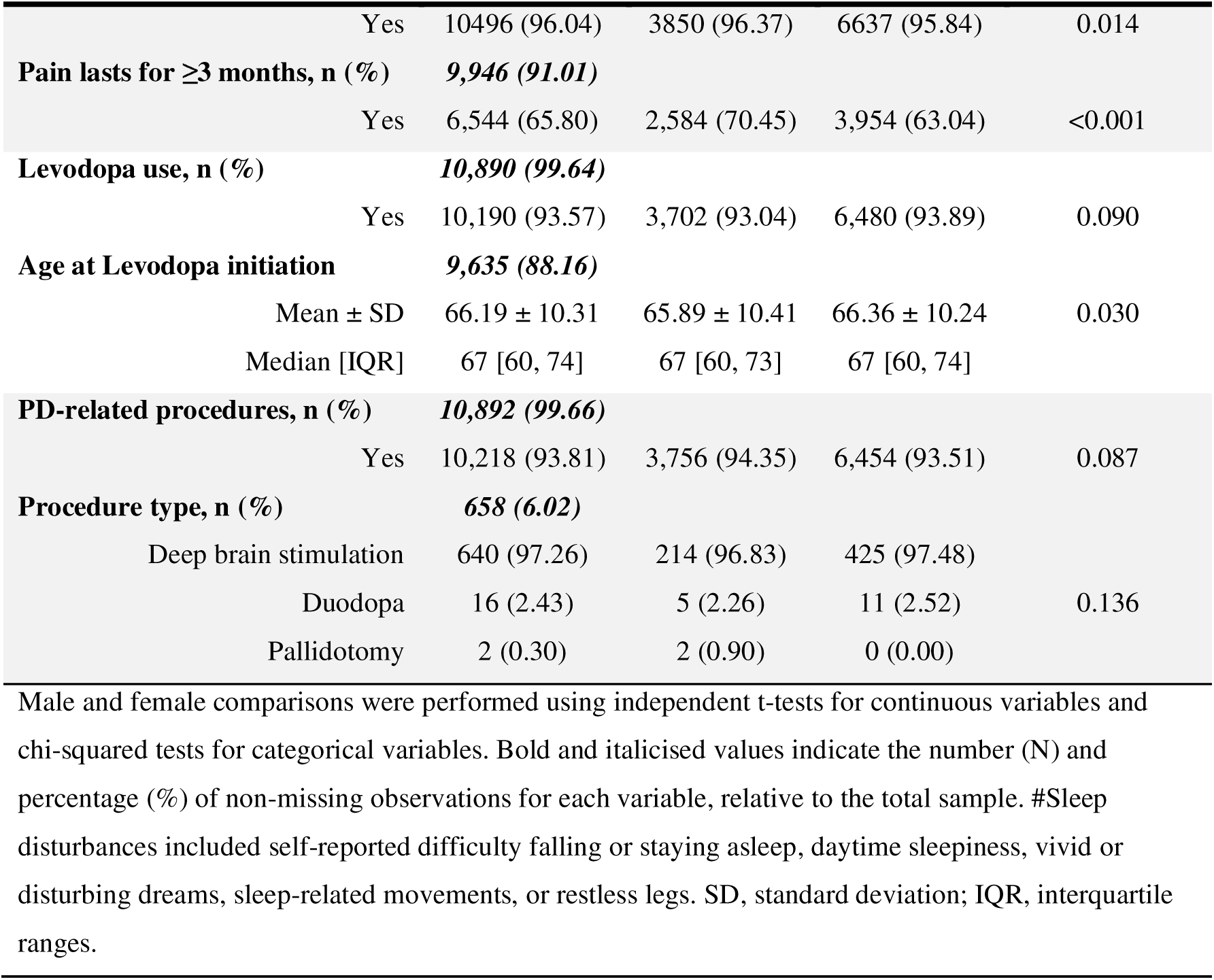
Parkinson’s disease-related outcomes in the APGS cohort.

Females had a slightly earlier symptom onset and were younger at both diagnosis and initiation of levodopa therapy, though differences were minimal (<1 year). A greater proportion of females experienced unilateral (81% vs. 75%) and left-sided onset (49% vs. 45%). Rigidity or stiffness at onset was more common among females (8% vs. 7%), while bradykinesia was more often reported by males (14% vs. 11%). Additionally, females were more likely to experience falls (45% vs. 41%) and pain (70% vs. 63%), whereas memory changes were more common among males (67% vs. 61%) (p = 0.03 to <0.001).

### Family history

The most commonly reported familial condition was PD, observed in 2,621 (24%) participants (Figure 2a). Among these, 38% reported affected parents, 22% siblings and 19% grandparents (Figure 2b). The majority (81%) had only one affected relative; eight participants reported five or more (Figure 2c). Dementia was the second most common familial condition (22%), followed by Alzheimer’s disease (15%) and tremor (6%).

**Figure 2.**
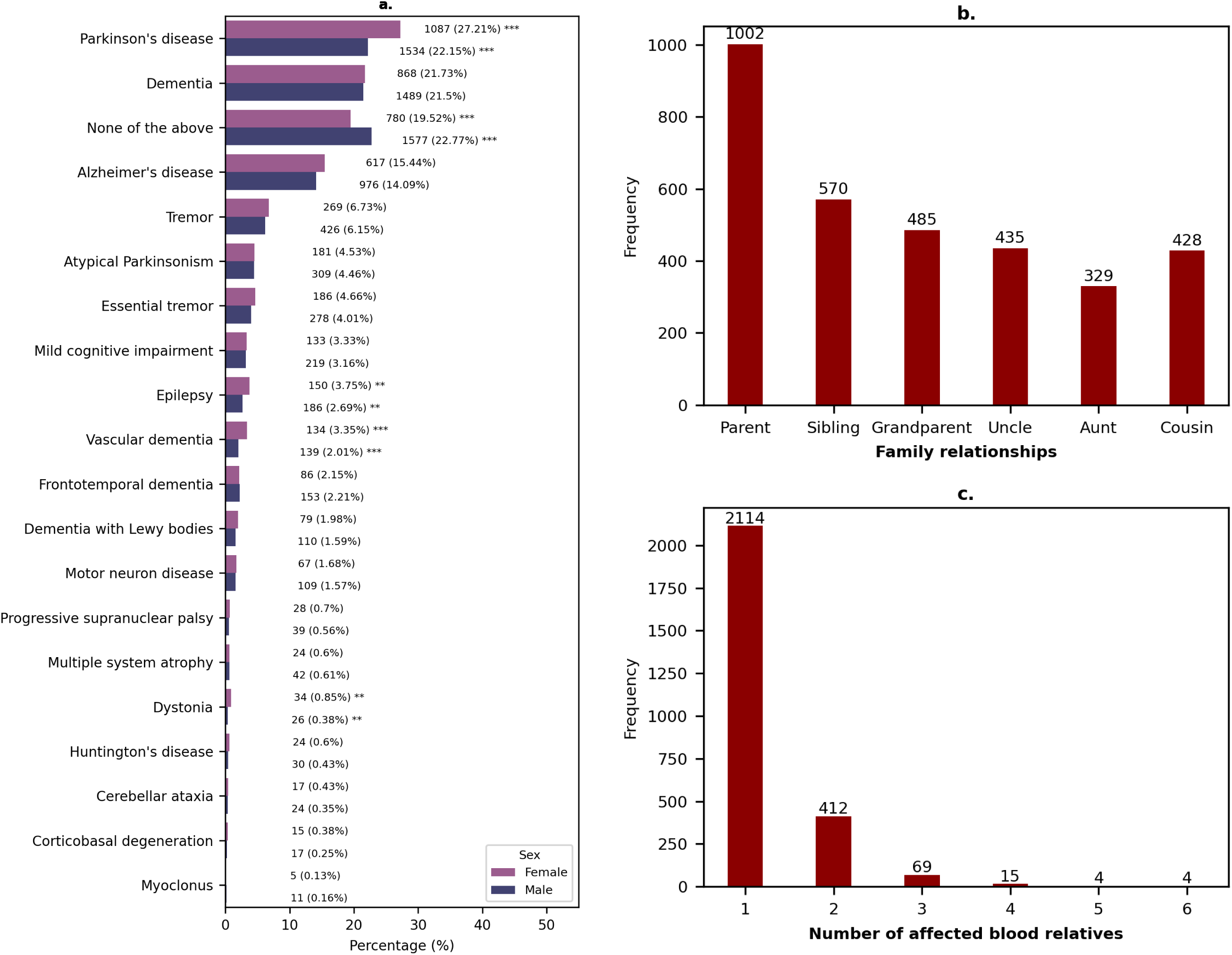
Family history by sex in the APGS cohort. **a.** Distribution of positive family history for various conditions, comparing males and females. Bars indicate counts with corresponding percentages. Statistical significance: *p < 0.05, **p < 0.01, ***p < 0.001. **b.** Types of blood relatives reported to have Parkinson’s disease (PD). **c**. Total number of blood relatives with PD reported by each participant.

Female participants more frequently reported a positive family history across all assessed conditions, with several disorders showing statistically significant differences. PD was reported by 27% of females vs. 22% of males. Similarly, reports of epilepsy (4% vs. 3%), vascular dementia (4% vs. 2%), and dystonia (0.9% vs. 0.4%) were more common among female participants (*p* = 0.002 to < 0.001).

### Lifestyle and environmental factors

Regular consumption of caffeinated beverages was common, with 76% reporting coffee intake and 70% tea intake, averaging two cups per day (Table 4). Alcohol use was also prevalent: 69% reported current alcohol consumption, and 31% indicated a history of heavy alcohol use. Regular well water consumption was reported by 5%. While active smoking was less widespread, second-hand smoke exposure was relatively common. Among those who had a history of smoking (38%), only 6% of them continued the smoking habit, while 62% reported exposure to second-hand smoke at home and 25% at work.

**Table 4.**
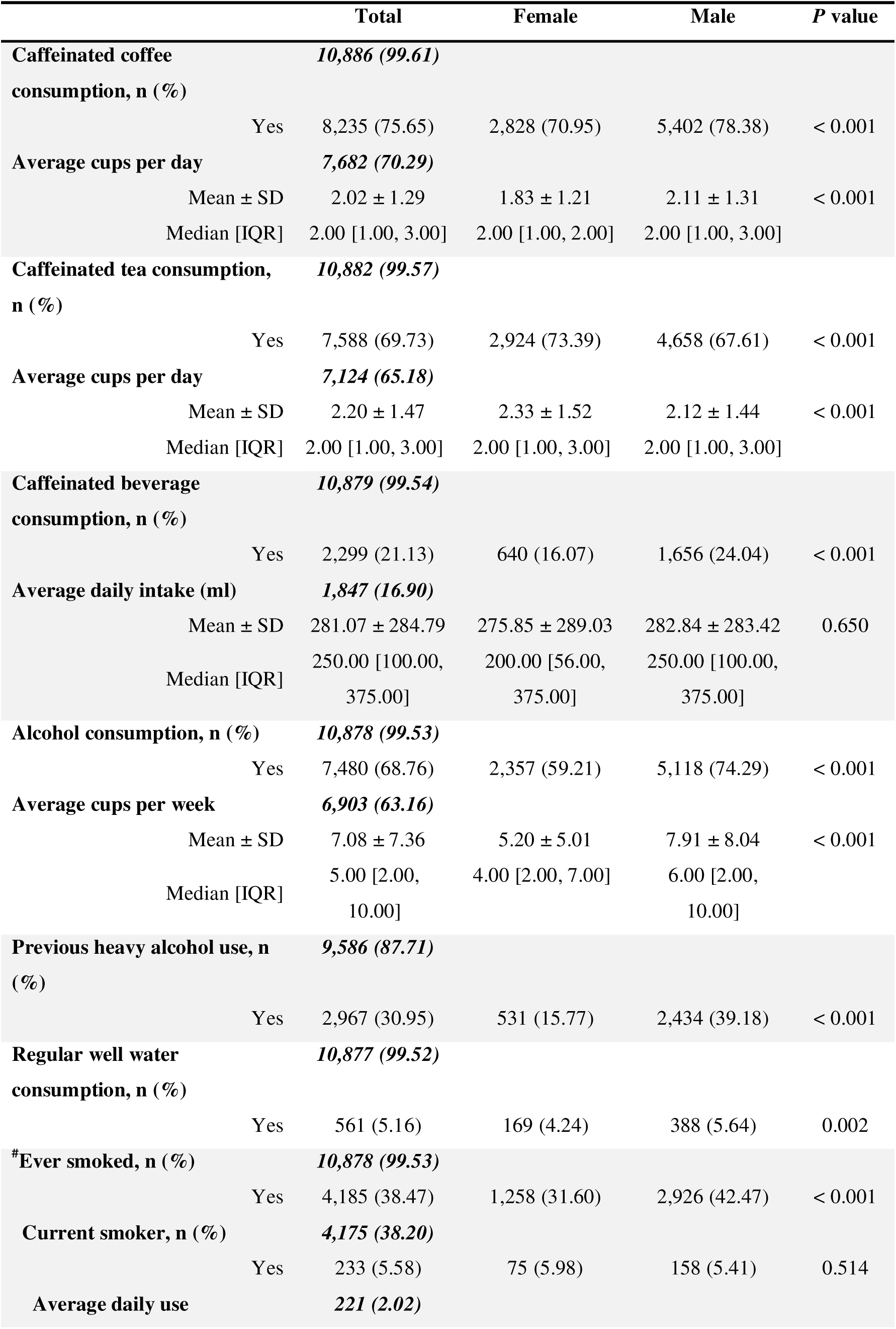
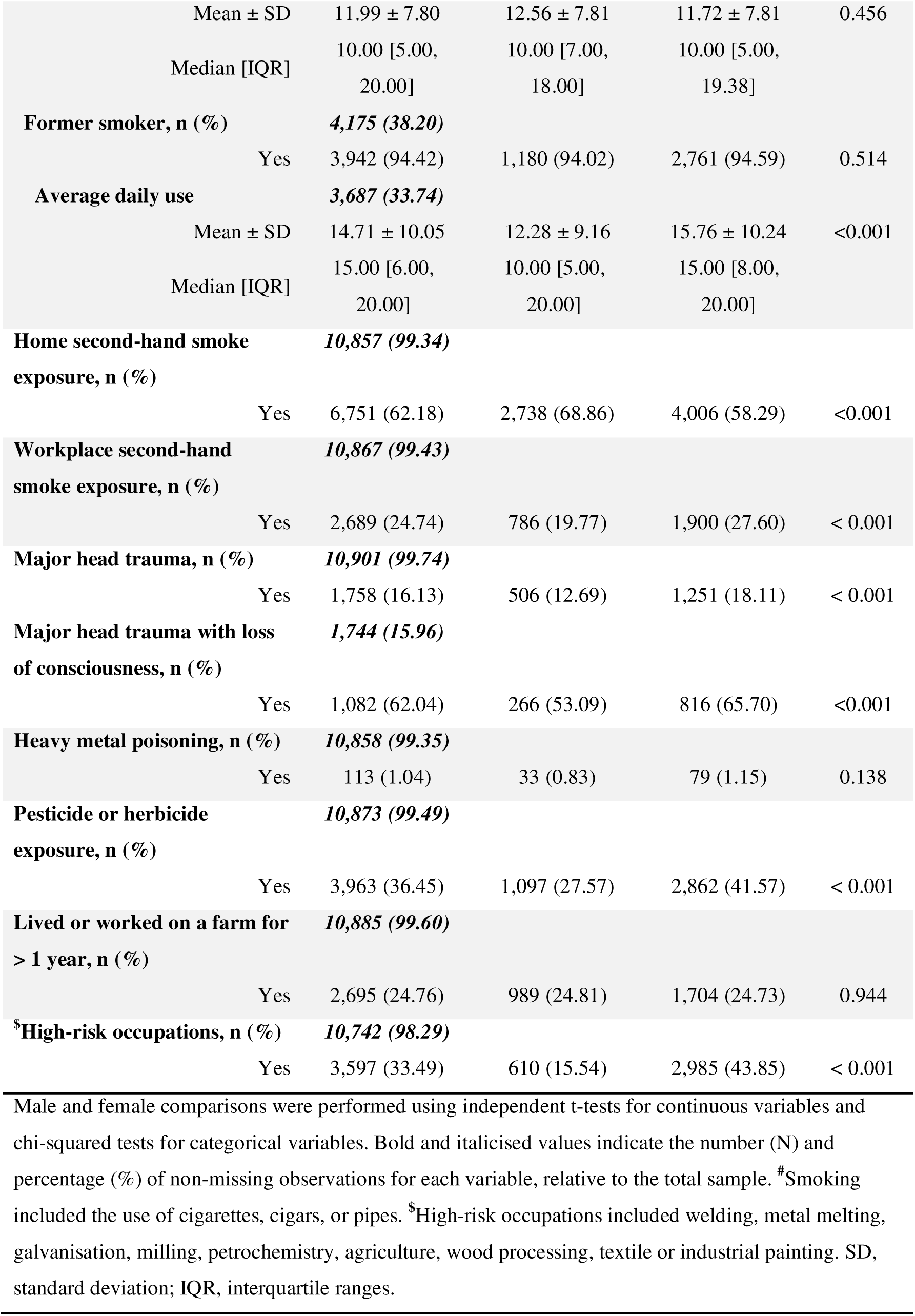
Lifestyle, events, and environmental risk factors in the APGS cohort.

Major head injury was reported by 16%, with 62% of these cases involving loss of consciousness. Pesticide or herbicide exposure was noted by 36%, and 25% had lived or worked on a farm for over one year. Additionally, 33% had worked in high-risk industries such as metal melting, galvanisation, milling, petrochemicals, agriculture, wood processing, textiles, or industrial painting.

Significant sex differences emerged across lifestyle and environmental variables. Males were more likely to consume coffee (78% vs. 71%), caffeinated beverages (24% vs. 16%), and alcohol (74% vs. 59%), both in prevalence and quantity. Well water consumption was also higher among males (6% vs. 4%). Conversely, females reported higher tea consumption (73% vs. 68%). A history of heavy alcohol use was reported by more than twice as many males as females (39% vs. 16%). While current and former smoking rates were comparable, a higher proportion of males had ever smoked (42% vs. 32%). Males also reported higher rates of major head injuries (18% vs. 13%), exposure to pesticides or herbicides (42% vs. 28%), and employment in high-risk occupations (44% vs. 16%) (*p* = 0.002 to <0.001).

### Medical history

Participants reported a broad range of co-occurring conditions. Common neurological and sleep-related disorders included restless legs syndrome (20%), migraine (20%), sleep apnoea (15%), essential tremor (12%), insomnia (11%), and rapid eye movement sleep behaviour disorder (RBD) (10%) (Figure 3). Mental health conditions were also prevalent: depression and anxiety affected 29% and 19% of participants, respectively.

**Figure 3.**
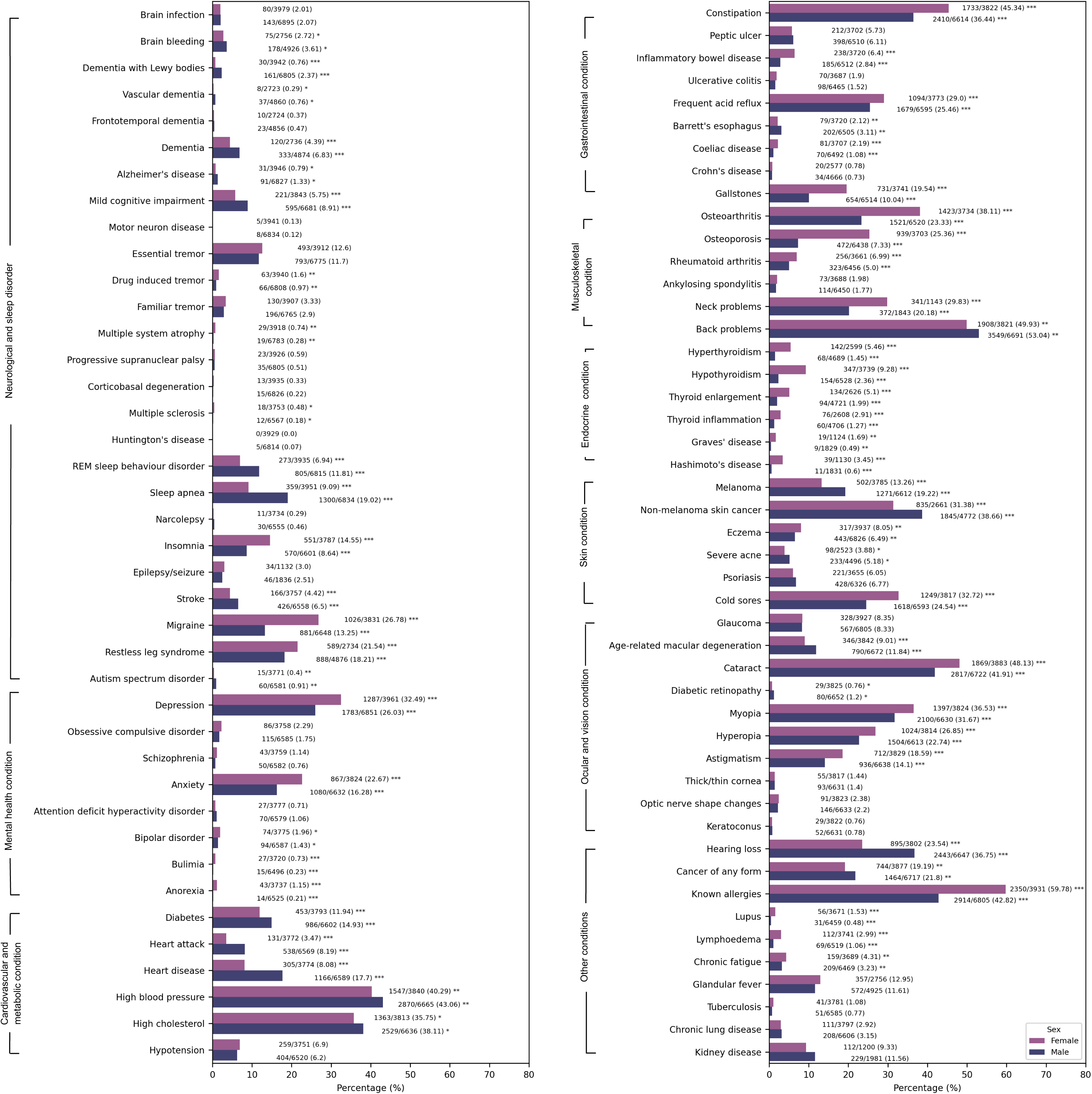
Comorbidities by sex in the APGS cohort. Labels display the number of affected participants, total respondents per item, and corresponding percentages. Statistical significance between male and female participants is indicated as: *p < 0.05, **p < 0.01, ***p < 0.001.

Among non-neurological comorbidities, back problems (52%), allergies (48%), cataracts (45%), and vision conditions (16%-34%) were frequently reported. Other common comorbidities included hypertension (41%), constipation (40%), high cholesterol (34%), cancers (17%-36%), hearing loss (30%), osteoarthritis (30%), cold sores (28%), and frequent acid reflux (26%).

Sex-based differences were evident across multiple domains. Females more often reported pain-related conditions, such as migraine (27% vs. 13%) and osteoarthritis (38% vs. 23%), as well as mental health conditions, including depression (32% vs. 26%) and anxiety (23% vs. 16%). In contrast, males had a higher prevalence of cardiovascular and metabolic conditions, including hypertension (43% vs. 40%), heart disease (18% vs. 8%), and diabetes (15% vs. 12%). Males also more frequently reported cognitive impairments, such as dementia (7% vs. 4%) and mild cognitive impairment (9% vs. 6%), along with sleep disorders including sleep apnoea (19% vs. 9%) and RBD (12% vs. 7%). Cancer diagnoses were also more frequent among males, ranging from 19% to 39%, compared to 13% to 31% in females (*p* = 0.042 to < 0.001).

### Impulsivity

Overall, most participants exhibited no or mild impulsivity. Impulsive behaviours like repetitive, mechanical tasks (punding) or hobbies (hobbyism), use of PD medication, eating, shopping, and sexual behaviours were more common than gambling (Figure 4).

**Figure 4.**
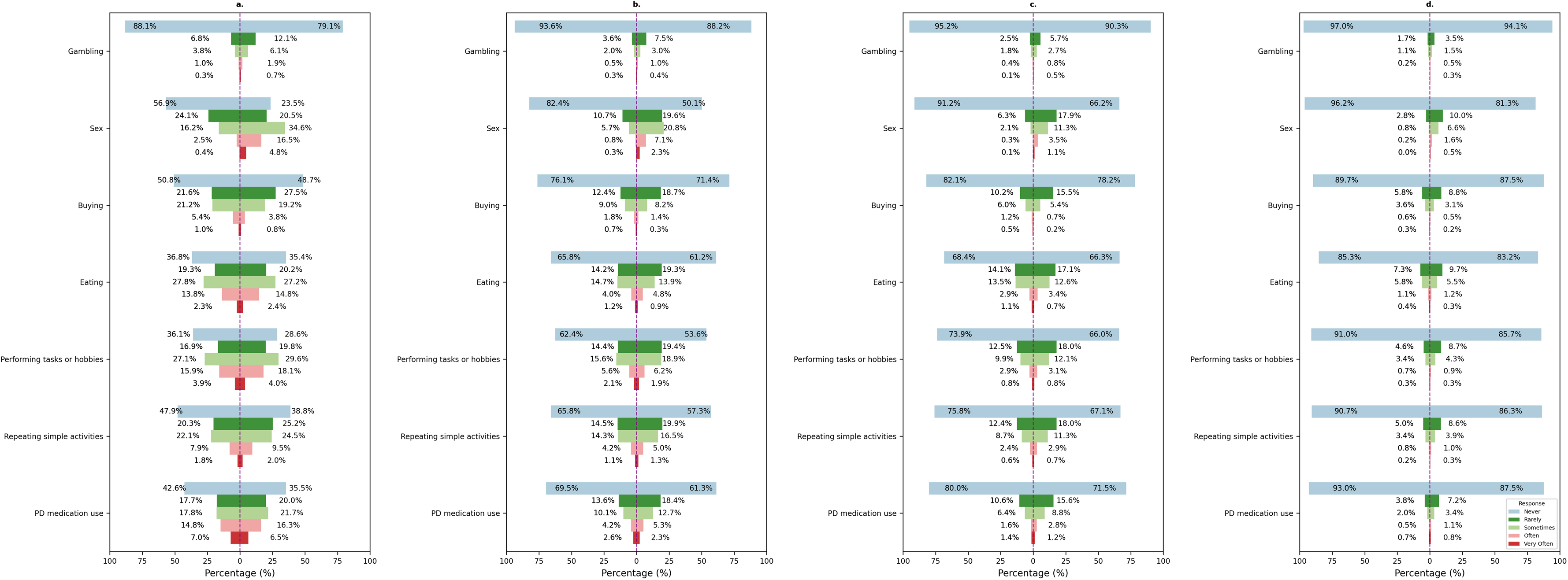
Impulse control behaviours by sex in the APGS cohort. This figure compares self-reported impulsive behaviours between female (left) and male (right) participants. Four questions assessed: **a.** frequency of intrusive thoughts or feelings of guilt related to specific behaviours; **b.** frequency of having urges or desires for these behaviours, including associated distress or irritability; **c.** frequency of having difficulties controlling the behaviours, such as escalation over time or inability to reduce them; and **d.** frequency of engaging in actions to sustain the behaviours, including concealment, deception, debt accumulation, or illegal activities. Significant differences between males and females were observed for all behaviours in Questions **a-d** (p = 0.001 for eating in Question **d** and p < 0.001 for all other behaviours), except for eating in Question **a** (p = 0.328).

Regarding control over these behaviours, up to 18% of participants reported “often” and 7% “very often” having difficulty controlling related thoughts (Figure 4a). In comparison, fewer than 10% experienced excessive urges that caused distress (Figure 4b), and under 4% struggled to control the behaviours themselves at regular cadence (Figure 4c). Actions to sustain these behaviours, like hiding, lying, accruing debt, or illegal activity, were rare (<7% reported doing so “sometimes”) (Figure 4d).

Sex differences were evident, with males reporting higher levels of impulsivity, particularly for sexual behaviour, punding, hobbyism, and gambling (*p* ≤ 0.001). For impulsive sexual behaviour, 56% of males vs. 20% of females reported difficulty suppressing intrusive thoughts, 31% vs. 7% experienced excessive urges, 16% vs. 3% struggled to control the behaviour, and 9% vs. 1% admitted to actions to sustain the behaviour.

COVID-19-related outcomes are detailed in the Supplementary Results (Supplementary Table S2).

## Discussion

The APGS (N=10,929) represents the largest nationwide PD cohort in Australia and the largest active PD cohort globally. Established through time- and cost-efficient recruitment strategies, APGS aims to uncover genetic and environmental factors influencing PD onset and progression, deepen understanding of underlying aetiological mechanisms, and ultimately inform the identification of novel therapeutic targets. Here, we presented a comprehensive characterisation of individuals with PD in APGS, covering a broad spectrum of sociodemographic, clinical, and environmental factors.

In APGS, the average participant age was 71 years, with a mean symptom onset at 64 and diagnosis at 68, consistent with previous reports showing a sharp rise in PD incidence beyond the age 60.^9,10^ A meta-analysis of 47 studies showed that global incidence increased from approximately 41 per 100,000 in the 40-49 age group to 428 per 100,000 in those aged 60-69 and 1,903 per 100,000 in individuals over 80.^9^ Ageing is a well-established PD risk factor. Although not solely responsible for disease onset, age-related declines in dopamine metabolism, protein degradation, and mitochondrial function heighten the vulnerability of *substantia nigra* neurons to genetic and environmental stressors.^12^

Male sex is another consistently reported risk factor, with studies across different populations showing that PD prevalence and incidence are roughly twice as high in men compared to women.^1,10,17^ This pattern is mirrored in APGS, where 63% of participants are male. One prevailing hypothesis posits that estrogen may exert neuroprotective effects on dopaminergic neurons, possibly through modulating nigrostriatal dopamine synthesis, metabolism, receptor binding, and anti-inflammatory pathways.^18^ However, sex-specific genetic mechanisms and environmental exposures may also contribute to the higher prevalence in males.^19,20^ In our study, male participants reported higher levels of education, greater alcohol consumption, higher smoking prevalence, and more frequent exposure to pesticides and high-risk occupations than females. Further research to disentangle the independent and interactive effects of sex, environment, and genetics is essential for informing the development of sex-specific treatment strategies.

Within our cohort, while tremor was the predominant initial symptom overall, females more often reported rigidity or joint stiffness, and males more frequently reported bradykinesia. Females also more frequently experienced falls and pain, whereas males reported cognitive impairment more often. Motor symptoms may appear later in females,^17,21^ possibly due to sex differences in striatal dopamine receptor expression in the striatum in women.^22^ Consistent with our findings, existing literature suggests that females with PD tend to experience more psychiatric symptoms, pain, autonomic dysfunction, postural instability, and higher risk of falls.^17,23^ In contrast, males appear more prone to earlier and more rapid cognitive decline, as well as sleep behaviour disorders (e.g., RBD).^17,21,23^ These observed sex differences may reflect distinct pathogenic pathways in males and females; however, mechanistic understanding of how biological sex influences disease processes and clinical progression remains limited and warrants further investigation.

Family history has been implicated as a key risk factor for PD, increasing the likelihood of developing the disease two-to threefold two-to threefold in both familial and sporadic forms.^24^ In APGS, one in four participants reported a family history of PD, reinforcing the significant role of genetic susceptibility. Notably, first-degree relatives, particularly parents and siblings, were most commonly reported, highlighting the influence of both inherited genetic variants and shared environmental exposures.^12,13^ While the majority reported only one affected relative, some described multiple family members, including rare familial clustering of five or more cases, which may reflect the presence of rare, highly penetrant variants. Beyond PD, family histories of other neurodegenerative disorders were also common among APGS participants, further supporting evidence of shared genetic risk and overlapping pathogenic pathways.^25,26^ Interestingly, female participants more frequently reported the aforementioned family histories, a pattern possibly explained by biological differences in disease expression, genetic susceptibility, or inheritance,^17,21^ as well as sociocultural factors such as greater engagement in family health matters.^27^

Environmental and lifestyle factors, including exposure to toxicants such as pesticides, solvents, heavy metals, and industrial pollutants has long been recognised as a risk factor, with specific pesticide effects replicated in animal models.^8,28^ Traumatic head injury is also associated with a higher risk and earlier onset of PD.^29^ Consistent with previous findings, a substantial proportion of APGS participants reported pesticide exposure and potential occupational contact with hazardous substances. Major head injury was reported by 16% of participants, markedly higher than the national estimate of 1 in 45.^30^ Conversely, only 6% were current smokers, compared to 11.1% in general Australian population.^30^ Lifestyle factors such as tobacco use, caffeine consumption, and diets rich in fruits, vegetables, and whole grains have been linked to reduced PD risk.^8,12^ However, evidence on these associations remains inconclusive,^1,8,10^ and further mechanistic research is needed for a more complete understanding of the underlying biological mechanisms.

The burden of comorbidities in PD is substantial, with some potentially involved in its pathogenesis. In the APGS cohort, we observed a high prevalence of gastrointestinal, cardiovascular, visual, and oncological conditions, while 10-29% reported psychiatric, neurological, and sleep disorders. Psychiatric comorbidities affect up to 89% of people with PD and are associated with worse prognosis and greater disability.^31,32^ The exact nature of these associations remains debated; these symptoms may stem from intrinsic disease processes, shared genetic or pathophysiological pathways, or psychological responses to diagnosis.^29,31^ Sleep disorders are also common in PD due to neurodegeneration in sleep-regulating brain regions,^33^ with some (e.g., RBD) preceding motor symptoms.^3,29^ The diverse comorbidities in PD highlight its clinical complexity, and understanding their molecular underpinnings may offer insights into biomarkers and targeted interventions.

Impulse control behaviours (ICBs) are a common complication of dopaminergic therapies in PD, particularly with dopamine agonists, and can have devastating psychosocial consequences on patients and caregivers.^34^ Prevalence estimates vary widely (14-60%),^31,34,35^ likely due to varying definitions and assessment tools. Notably, a 17.5% prevalence reported prior to treatment also suggests a possible predisposition in some individuals.^35^ In APGS, although most cases were mild, 25% of participants experienced difficulties controlling impulsive thoughts at regular cadence. Males exhibited greater impulsivity, especially in relation to sexual behaviour. This aligns with existing literature linking male gender to higher impulsivity and hypersexuality, potentially driven by neurobiological factors such as stronger mesocorticolimbic reward circuitry, as well as psychological and sociocultural influences. ^21,35^ These findings reinforce the high prevalence and clinical significance of ICBs in PD, underscoring the need for early identification and appropriate management.

Our study has several limitations. First, the response rate was modest (∼10,000 participants from 186,000 invitations), raising the possibility of self-selection bias. Second, while the remote design enabled national reach, it may compromise data accuracy and limit the depth of clinical phenotyping. Future study phases will incorporate digital biomarkers from smartphones and wearables, alongside electronic patient-reported outcomes, to support objective, longitudinal, and patient-centred data collection. Third, the cohort is predominantly of European ancestry, which may limit the generalisability of findings to more diverse populations. Lastly, the findings are limited by the study’s descriptive nature and absence of an age-and sex-matched control group, with recruitment still underway.

## Conclusions

As the largest active PD cohort, the APGS offers an unprecedented opportunity to uncover the sociodemographic, genetic, and environmental drivers of disease susceptibility, presentation, and progression. Here, we provided a comprehensive profile of the PD cohort, forming a critical foundation for transformative research. By contributing to national and global efforts, APGS is poised to expedite the discovery of novel biomarkers and therapeutic targets, ultimately accelerating the development of strategies to prevent, slow, or modify the course of PD.

## Data Availability

An anonymised version of the data used in this study may be made available to qualified researchers who submit a collaboration proposal to the APGS investigators, are able to sign an institutional data transfer agreement, and agree to comply with all conditions intended to protect participant privacy and confidentiality.

## Contributors

F.Cao performed all statistical analyses and wrote the first draft of the manuscript. K.M., R.P., M.F., R.A.C., and S.E.L.-A. were responsible for study operations, including participant engagement, saliva sample collection and processing, and implementation of the electronic questionnaire, among other tasks. N.S.O., L.M.G.-M., S.D.-T., V.F.-O., Z.C., and F.Chafota contributed to the interpretation of data and provided input on the analysis and manuscript structure. V.M. and C.C. actively contributed to participant recruitment through media engagement and advocacy. C.M.S. and K.R.K. contributed clinical expertise and reviewed the manuscript for accuracy in clinical interpretation. G.D.M. and N.G.M. helped design the study, including the questionnaire, and provided senior scientific guidance. M.E.R. conceived and led the study, coordinated the research team, and oversaw the preparation and revision of the manuscript. All authors critically reviewed the manuscript for intellectual content and approved the final version for submission.

## Data sharing

The data supporting the findings of this study are not publicly accessible due to participant privacy considerations. However, anonymised datasets can be made available to qualified researchers upon request to the corresponding author. Data sharing will be contingent upon obtaining all necessary ethical approvals and signing an institutional data transfer agreement. Researchers may contact the corresponding author to request access to the code used for data processing and analysis.

## Acknowledgements

We are deeply grateful to all the participants of the Australian Parkinson’s Genetics Study (APGS) whose invaluable contributions and commitment made this research possible. We would like to thank Michael Katz and Richard Balanson for their valuable input and contributions in setting up the study. We thank the Health Data Analysis and Strategy Branch team at Services Australia for their kind assistance and facilitating the mail-outs. APGS is supported by the Shake It Up Australia Foundation and the Michael J. Fox Foundation for Parkinson’s Research (MJFF-021952). MER thanks the support from the Rebecca L. Cooper Medical Research Foundation (F20231230). KRK is supported by grants from the Medical Research Future Fund, the Lord Mayor’s Charitable Fund, and the Ainsworth 4 Foundation (unrelated to the current study).

