## Supplementary Materials for "Insights from a nationwide study of 10,929 Australians living with Parkinson’s disease: Risk factors, comorbidities, and sex differences"

### Supplementary Methods

#### Recruitment

The Australian Parkinson's Genetics Study (APGS) collaborates with *Services Australia* to implement a large-scale, assisted mail-out to individuals across the country who have been prescribed Parkinson's disease (PD)-related medications. *Services Australia* is a government agency that delivers a range of social and health-related services to Australian citizens and permanent residents. Among its responsibilities, the agency administers the Pharmaceutical Benefits Scheme (PBS), a national program that subsidises prescription medicines. (Further details and a full list of subsidised medicines can be found at https://www.pbs.gov.au/pbs/). The agency maintains prescription records from the past five years and, with appropriate ethics approval, supports health research by coordinating mail-outs on behalf of research organisations. Importantly, the recipient's personal data remain secure and confidential throughout this process.

Following a successful pilot phase, and with the support of Global Parkinson's Genetics Program (GP2), the Shake It Up Australia Foundation, and the Michael J. Fox Foundation, full-scale nationwide recruitment was launched in April 2022. The rollout began during *Parkinson's Awareness Month*, accompanied by a national media campaign and public appeal. In September 2022, 100,000 invitation letters were distributed by *Services Australia* to individuals who met the eligibility criteria. (See Supplementary Table S1 for a list of included medications and their corresponding PBS codes). Subsequent recruitment waves followed with 60,000 additional letters distributed in July 2023, and a further 26,000 letters in June 2024.

Control recruitment began in February 2023 through a mailout sent to participants of the QSkin Sun and Health Study,[1] and in 2024, individuals within the APGS cohort were approached and asked to refer non-blood relatives as controls for the study. The eligibility criteria for controls were: 1) aged 45 years or older, 2) no family history or diagnosis of PD, and 3) residing in Australia at the time of participation.

#### Saliva sample collection

Upon completion of the questionnaire, participants who met the eligibility criteria were invited to provide a saliva sample for genetic analysis. Those who consented to participate in the genetic component of the study were contacted by the research team to confirm their details and mailing address. A saliva collection kit (Isohelix or Thermo Fisher), along with detailed instructions for sample collection and a prepaid return envelope, was then dispatched to the participant's nominated address. Participants were asked to self-collect their saliva sample and return it by traditional post to our research laboratory in Brisbane for processing. Most participants have been genotyped, with a subset undergoing whole-genome sequencing from saliva samples.

#### Sub-studies

Several focused sub-study initiatives are supported within the APGS platform to enhance its translational potential, enabling future discovery, targeted interventions, and the advancement of precision medicine in PD. These include:

- MonoPDAus[2] - Investigates monogenic forms to improve gene discovery and genetic diagnosis.
- Cognition and PD - Examines cognitive-related aspects of PD.
- Speech and Parkinson's - Focuses on characterising speech and language changes to better understand symptomatology and identify diagnostic and prognostic biomarkers.

### Supplementary Table S1 Anti-Parkinson's disease medication name and corresponding PBS code

| Medication | PBS code |
|---|---|
| Levodopa + carbidopa | 1242J, 1245M, 1255C, 8970D, 9743T, 9744W |
| Levodopa + benserazide | 2225D, 2226E, 2227F, 2228G, 2229H, 2231K, 8218M, 8219N |
| Levodopa + carbidopa + entacapone | 8797B, 8798C, 8799D, 9292C, 9344T, 9345W |
| Amantadine hydrochloride | 3016R |
| Apomorphine hydrochloride hemihydrate | 10950H, 10971K, 11083H, 11093W, 5609F, 5610G, 9607P, 9640J |
| Cabergoline | 8393R, 8394T |
| Pramipexole dihydrochloride monohydrate | 3418X, 3419Y, 3420B, 3421C, 3422D, 5143Q, 5145T, 9151P, 9152Q, 9153R, 9393J, 9394K |
| Rotigotine | 1140H, 2384L, 2385M, 2410W |
| Benztropine mesilate | 11249C, 11255J, 11265X, 2362H |
| Trihexyphenidyl (benzhexol) hydrochloride | 1109J, 1110K |
| Rasagiline | 1952R |

### Supplementary Results

#### Supplementary Table S2 COVID-19 Vaccination, Infection History, and Clinical Outcomes in the APGS Cohort

| | All | Female | Male | *P* value |
|---|---|---|---|---|
| **Vaccination, n (%)** | ***7,509 (68.71)*** | | | |
| Yes | 7,398 (98.52) | 2,652 (98.88) | 4,742 (98.34) | 0.077 |
| **Vaccination dose, n (%)** | ***7,395 (67.66)*** | | | |
| Four or more | 5,469 (73.96) | 1,955 (73.63) | 3,511 (74.13) | 0.744 |
| Three | 1,490 (20.15) | 536 (20.19) | 954 (20.14) | |
| Two | 389 (5.26) | 149 (5.61) | 240 (5.07) | |
| One | 47 (0.64) | 15 (0.56) | 31 (0.65) | |
| **Positive history, n (%)** | ***7,500 (68.62)*** | | | |
| Yes | 3,340 (44.53) | 1,157 (43.14) | 2,181 (45.31) | 0.073 |
| **Number of infections, n (%)** | ***3,241 (29.66)*** | | | |
| Mean ± SD | 1.16 ± 1.78 | 1.14 ± 0.40 | 1.18 ± 2.18 | 0.394 |
| **Contraction timing, n (%)** | ***3,278 (29.99)*** | | | |
| Post-vaccination | 2,823 (86.12) | 978 (85.86) | 1,844 (86.29) | 0.778 |
| Pre-vaccination | 455 (13.88) | 161 (14.14) | 293 (13.71) | |
| **Symptom severity, n (%)** | ***3,320 (30.38)*** | | | |
| Mild | 1,846 (55.60) | 633 (55.00) | 1,211 (55.88) | 0.051 |
| Moderate | 1,045 (31.48) | 383 (33.28) | 662 (30.55) | |
| Severe | 236 (7.11) | 84 (7.30) | 152 (7.01) | |
| No symptoms at all | 193 (5.81) | 51 (4.43) | 142 (6.55) | |
| **Hospitalisation, n (%)** | ***3,317 (30.35)*** | | | |
| Yes | 129 (3.89) | 36 (3.13) | 93 (4.30) | 0.120 |

Male and female comparisons were performed using independent t-tests for continuous variables and chi-squared tests for categorical variables. Bold and italicised values indicate the number (N) and percentage (%) of non-missing observations for each variable, relative to the total sample. SD, standard deviation; IQR, interquartile ranges.
